# Critical care capacity in Africa: Post-pandemic ICU capacity, service readiness, and patient profiles across public and private hospitals in Ethiopia

**DOI:** 10.1101/2025.07.04.25330893

**Authors:** Tesfay Yohannes, Fitsum K Belachew, Degisew Derso, Selam Daniel, Azeb Demelash, Kalkidan Kifle, Wagari Tuli Nora, Elubabor Buno Teko, Emnet Tesfaye, Yared Boru, Rupert Pearse, Menbeu Sultan, ENCCA collaborators

## Abstract

**Background:** The COVID-19 pandemic highlighted critical care disparities in low-resource settings. Ethiopia implemented interventions to strengthen ICU capacity post-pandemic, yet gaps persist. This study evaluates national ICU capacity post-COVID-19, including private facilities, and identifies challenges for equitable critical care delivery.

**Methods:** A cross-sectional nationwide assessment was conducted in public and private hospitals across Ethiopia. Data were collected through site visits and standardized questionnaires, assessing ICU capacity, staffing, equipment, protocols, and emergency preparedness. Patient-level data were also collected to assess common admission characteristics. Findings were compared with pre-COVID-19 baseline data to evaluate progress and identify gaps following the implemented interventions.

**Findings:** A total of 159 hospitals across Ethiopia were surveyed, achieving a 93% response rate, with 117 out of 159 (73·58%) being governmental and 42 out of 159 (26·42%) being private facilities, assessing a total of 1028 beds. Post-COVID-19, public ICU facilities increased from 51 to 117, with beds increasing from 324 to 762. Private facilities contributed 266 beds (25.9% of national capacity). Improvements included 24/7 ICU-trained physician availability (52.1% vs. 29.0% pre-COVID) and disaster preparedness plans (21.4% vs. 6.0%). Persistent gaps included advanced hemodynamic monitoring (5/117 public, 3/42 private facilities) and organ support (9/117 public ICUs). Among 279 admissions (mean age, 39.1 years; 55.2% male), neurological (32.1%) and respiratory (25.8%) conditions predominated, with sepsis accounting for 29.4% of cases. Hypertension (25.1%) and diabetes (17.2%) were the most common comorbidities.

**Interpretation:** This study reveals significant growth in Ethiopia’s ICU infrastructure and workforce following the COVID-19 pandemic, with a threefold increase in ICU beds and improved distribution. However, gaps in advanced monitoring, organ support, and referral coordination reveal systemic issues in critical care readiness. The high incidence of sepsis among mostly young patients underscores the need for targeted investment in essential emergencies and critical care (EECC) at all facility levels. Strengthening public-private integration, standardizing referral protocols, and scaling low-cost, high-impact interventions are crucial for enhancing equity and outcomes in resource-limited settings.

## Background

Critical care, which includes timely interventions for patients with life-threatening yet potentially reversible conditions, is a vital part of health systems (1,2). Intensive Care Units (ICUs) play a central role by delivering continuous monitoring and advanced organ support for critically ill patients (3). However, ICU capacity varies widely across countries. While some low- and middle-income countries (LMICs), such as South Sudan and Nauru, have no ICU beds, others like Kazakhstan report 21·3 beds per 100,000 population (4,5). In Africa, ICU bed availability remains low, with only Egypt, South Africa, and Seychelles exceeding 5 beds per 100,000 population (4,5).

LMICs face a disproportionate burden of critical illness and markedly higher mortality rates compared to high-income countries (6,7). These disparities, highlighted further by the COVID-19 pandemic, are driven by limited ICU infrastructure, shortages of trained personnel, and fragmented referral systems (8–10). Resource constraints contribute not only to insufficient critical care delivery but also to delayed recognition and treatment of patient deterioration, factors that compound poor outcomes, especially in sub-Saharan Africa (11–13).

In Ethiopia, a 2020 national review revealed substantial gaps in ICU services, including limited beds, lack of equipment, and workforce shortages (14). Moreover, the assessment focused only on public-sector ICUs and did not capture private facilities or patient-level data (14). In response, the Ethiopian Ministry of Health, along with its partners, launched several initiatives to expand ICU capacity, improve oxygen access, and strengthen workforce training (15,16). However, the national impact of these interventions remains unquantified.

This study addresses that gap by conducting a nationwide cross-sectional assessment of ICU capacity and patient case mix in public and private hospitals across Ethiopia. By capturing both facility- and patient-level data, this analysis provides a comprehensive view of Ethiopia’s critical care landscape and offers insights relevant to scaling up essential emergency and critical care services in other low-resource settings.

## Method

### Study design

This study used a nationwide, cross-sectional study design to assess ICU services across Ethiopia. Data collection took place from March to October 2024 and tried to involve all public and private healthcare facilities with operational ICUs.

### Study tool design

The survey tool was adapted from the guidelines of the World Federation of Intensive Care Medicine (WFICC) and a previous survey conducted in 2019. It was further refined to address limitations identified in earlier data collection rounds(3,14). The survey tool consisted of four forms, designed to evaluate ICU practices and patient care comprehensively. These forms assessed key areas critical to understanding ICU operations and their capacity to deliver care:

- ICU infrastructure: design, space utilization, and layout.
- Staffing: availability and qualifications of physicians, nurses, and allied health professionals, including nurse-to-patient ratios.
- Resources and equipment: monitoring capabilities and support for organ function.
- Educational and training opportunities: presence of formal professional development programs and the ICU’s role as a training center.
- Outreach and integration: ICU services linked to emergency departments, hospital wards, and follow-up care for discharged patients.
- Research and quality improvement capacity: ICU care providers’ involvement in advancing care standards and outcomes via research and quality improvement
- Scalability: Ability to respond and expand during disasters

### Each of the survey forms was tailored to address these areas systematically

- General information form: collected hospital-level data, including hospital type, total number of available beds, and the number of ICUs.
- ICU-Specific form: captured detailed information about ICU operations, including staffing levels, equipment availability, and the presence of standardized protocols. For hospitals with multiple ICUs, this form was completed for each unit individually.
- ICU observation form: focused on direct observation of one selected ICU per hospital. This form recorded data on cleanliness, organization, care practices, patient monitoring frequency, and adherence to infection control measures.
- Patient and admission assessment form: collected patient-level data, including demographics and diagnoses, from the medical records of patients admitted on the day of the visit. However, no patient identifiers, such as names, phone numbers, or other sensitive information, have been collected.

### Data Collection Preparation

A formal request was made to regional health administrators to recruit qualified data collectors with experience in ICU settings, such as emergency and critical care physicians, emergency and critical care nurses, anesthesiologists, and general practitioners. These individuals were assigned to conduct surveys in ICU facilities within their respective regions but were restricted from surveying hospitals where they were employed to avoid potential biases.

A one-day training session was conducted for all data collectors and supervisors. During the training, participants were oriented on data collection procedures, familiarized with survey variables, and provided with a Standard Operating Procedure (SOP) document. The session included discussions to clarify each variable, ensuring consistency in data collection. To facilitate real-time communication and support, a dedicated Telegram group was created for all participants, easing communications and ensuring data quality throughout the process. The study coordinators further supervised data collection to ensure adherence to protocols, address challenges, and provide ongoing support.

### Data Quality Control

Data entry and quality monitoring were conducted through a dedicated digital data collection platform, specifically designed for this study. Each regional and city administration health bureau focal person was granted access to oversee data entry for their respective regions. They ensured data completeness and accuracy by regularly reviewing submissions, while a national technical working group provided overarching supervision to ensure adherence to data collection protocols and maintain high-quality standards.

### Statistical Analysis

Data were exported from the dedicated digital data collection platform to Excel and analyzed using STATA version 17. Descriptive statistics were employed to summarize facility characteristics, while comparative analyses were conducted to compare results from the pre-COVID-19 study to the current post-COVID-19 status. Comprehensive reports were generated for each hospital and each region, with results meticulously validated through cross-checks with regional health bureau representatives to ensure precision and reliability.

### Ethical approval

Ethical approval was obtained from the Institutional Review Board (IRB) of Hawassa University College of Medicine and Health Sciences (protocol number IRB/333/16). The IRB waived the requirement for written informed consent, as the study was observational in nature. The Ministry of Health (MOH), a key collaborator in the project, facilitated communication with regional health offices through official letters and ensured their cooperation during the data collection process.

## Results

### General overview

As shown in **Figure 1**, this study assessed a total of 159 hospitals from all 14 regions and city administrations in Ethiopia, achieving a response rate of 93%. Twelve hospitals from Oromia Regional State were not included due to instability in the region. Of the surveyed hospitals, 117/159 (73·6%) were governmental, while 42/159 (26·4%) were private facilities. A total of 190 ICUs were assessed, with a combined bed capacity of 1028 beds, ranging from 2 to 37 beds per ICU. The distribution of ICUs per facility varied, with 136 having one ICU, 17 having two, 4 having three, and 2 having four.

**Figure 1:**
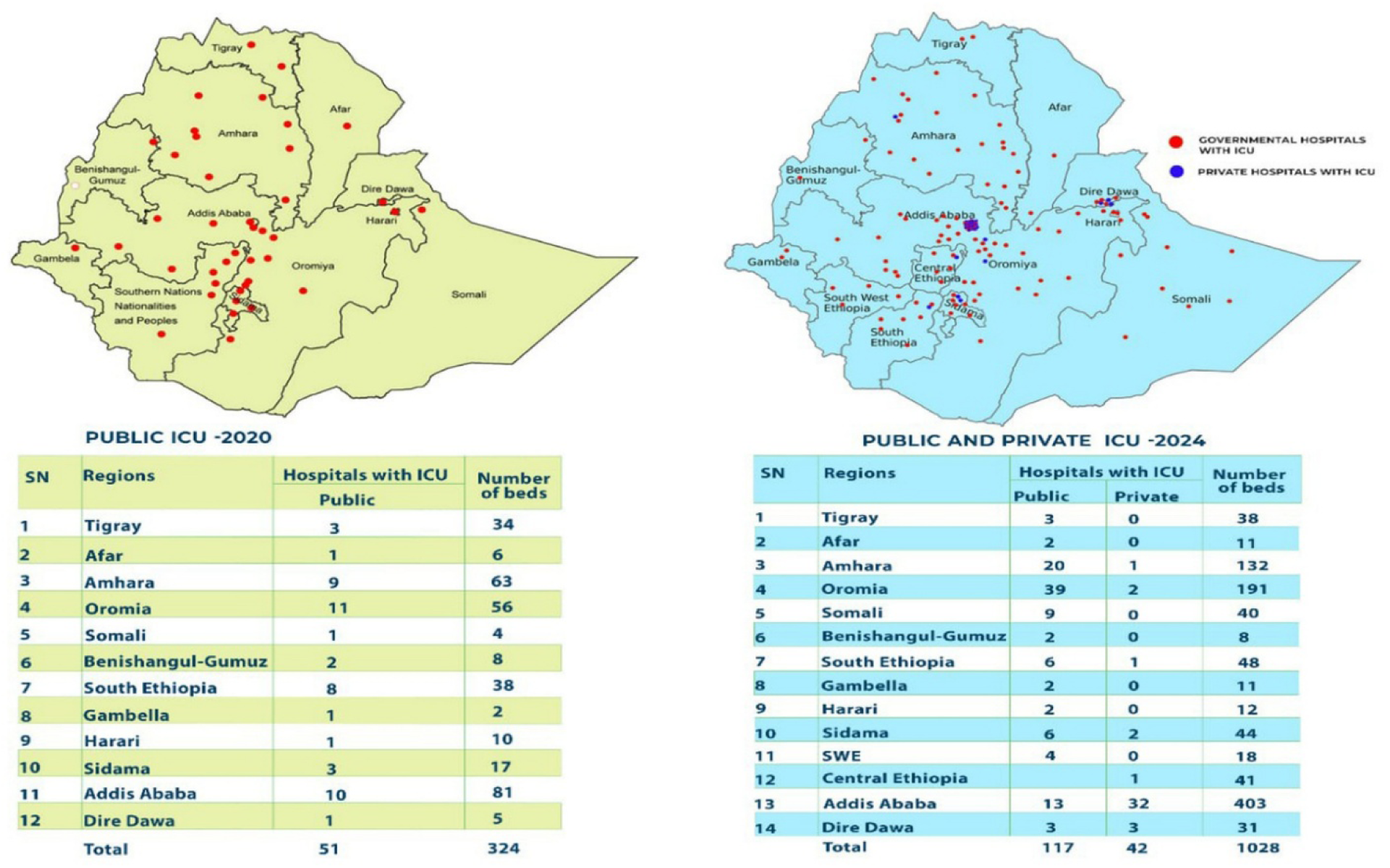
Distribution of ICUs Across Regions Before and After COVID-19.

### Comparison of ICU and Bed Capacity Pre- and Post-COVID-19

As shown in **Figure 1- Map and Table 1**, there has been a substantial increase in ICU facilities across Ethiopia after COVID-19, with hospitals with ICUs rising from 51 to 117 and total ICU beds increasing from 324 to 762, excluding private. Addis Ababa and Oromia saw the largest expansions, with ICU beds increasing from 81 to 186 and 56 to 181, respectively. The Somali region showed significant improvement, with hospitals equipped with ICUs increasing from 1 to 9 and beds rising from 4 to 40. Meanwhile, Sidama doubled its number of hospitals with ICUs, from 3 to 6, and beds increased from 17 to 31. The regions previously grouped under SNNPR, now subdivided into three regions (Southwest, Central, and South Ethiopia), collectively expanded from 8 hospitals with ICUs and 38 beds to 14 hospitals with ICUs and 116 beds. Conversely, regions like Benishangul-Gumuz showed no change, maintaining 2 hospitals with ICUs and 8 beds.

**Table 1:** Current Regional Distribution of ICUs, and ICU Beds.

| Regions | Hospitals with ICU |  | Total number of ICUs | Total beds (range/ICU) | Number of patients in the survey |
| --- | --- | --- | --- | --- | --- |
|  | Governmental | Private |  |  |  |
| Addis Abeba | 13 | 32 | 45 | 403(2-37) | 79 |
| Oromia | 39 | 2 | 41 | 191(2-24) | 64 |
| Amhara | 20 | 1 | 21 | 132(2-26) | 45 |
| Southwest Ethiopia | 4 | 0 | 4 | 18(4-6) | 11 |
| Central Ethiopia | 6 | 1 | 7 | 41(2-11) | 10 |
| South Ethiopia | 6 | 1 | 7 | 48(5-12) | 15 |
| Tigray | 3 | 0 | 3 | 38(6-24) | 7 |
| Sidama | 6 | 2 | 8 | 44(4-10) | 15 |
| Benishangul-Gumuz | 2 | 0 | 2 | 8(4) | 3 |
| Afar | 2 | 0 | 2 | 11(4-7) | 4 |
| Dire Dawa | 3 | 3 | 6 | 31(3-7) | 10 |
| Gambella | 2 | 0 | 2 | 11(5-6) | 4 |
| Harari | 2 | 0 | 2 | 12(4-8) | 4 |
| Somali | 9 | 0 | 9 | 40(2-11) | 8 |
| **Total** | **117** | **42** | **159** | **1028(2-37)** | **279** |

**Abbreviations:** ICU, intensive care unit.

### Comparison of Service Levels Pre- and Post-COVID-19

The comparison across the 12 domains highlights significant improvements in ICU capacity and services post-COVID-19. Facilities with 24/7 ICU-trained physicians increased from 15/51(29%) to 61/117(52·1%), and advanced organ support resources, such as CRRT, were introduced in 9/117(7·7%) (9 facilities) of surveyed ICUs. The number of ICUs rose from 78% (40 facilities) to 98·3% (115 facilities), and disaster preparedness plans increased from 3/51(6%) to 25/117(21·4%). Although some percentages declined, such as noninvasive monitoring (73% to 64·96%) and referral roles (61% to 32·48%), the actual number of facilities offering these services increased significantly. However, areas requiring further development include the availability of advanced organ support (limited to 9 facilities), nurse-to-patient ratios (50 facilities not meeting a <1:2 ratio), regular educational engagement (9 facilities offering consistent programs), and advanced hemodynamic monitoring, which remains available in only 5 facilities (4·27%), as further shown in **Tables 2 and 3**.

**Table 2:**
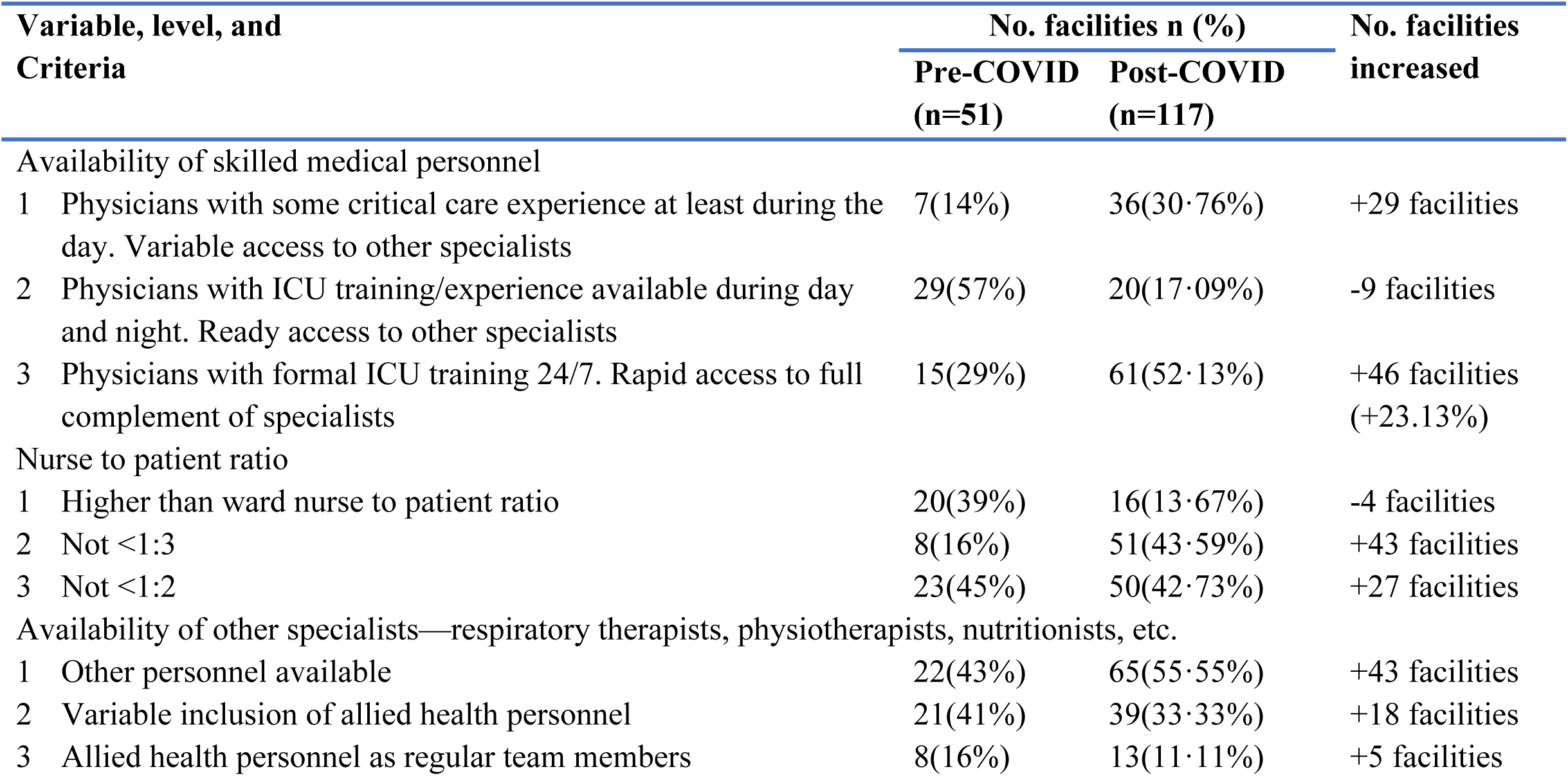
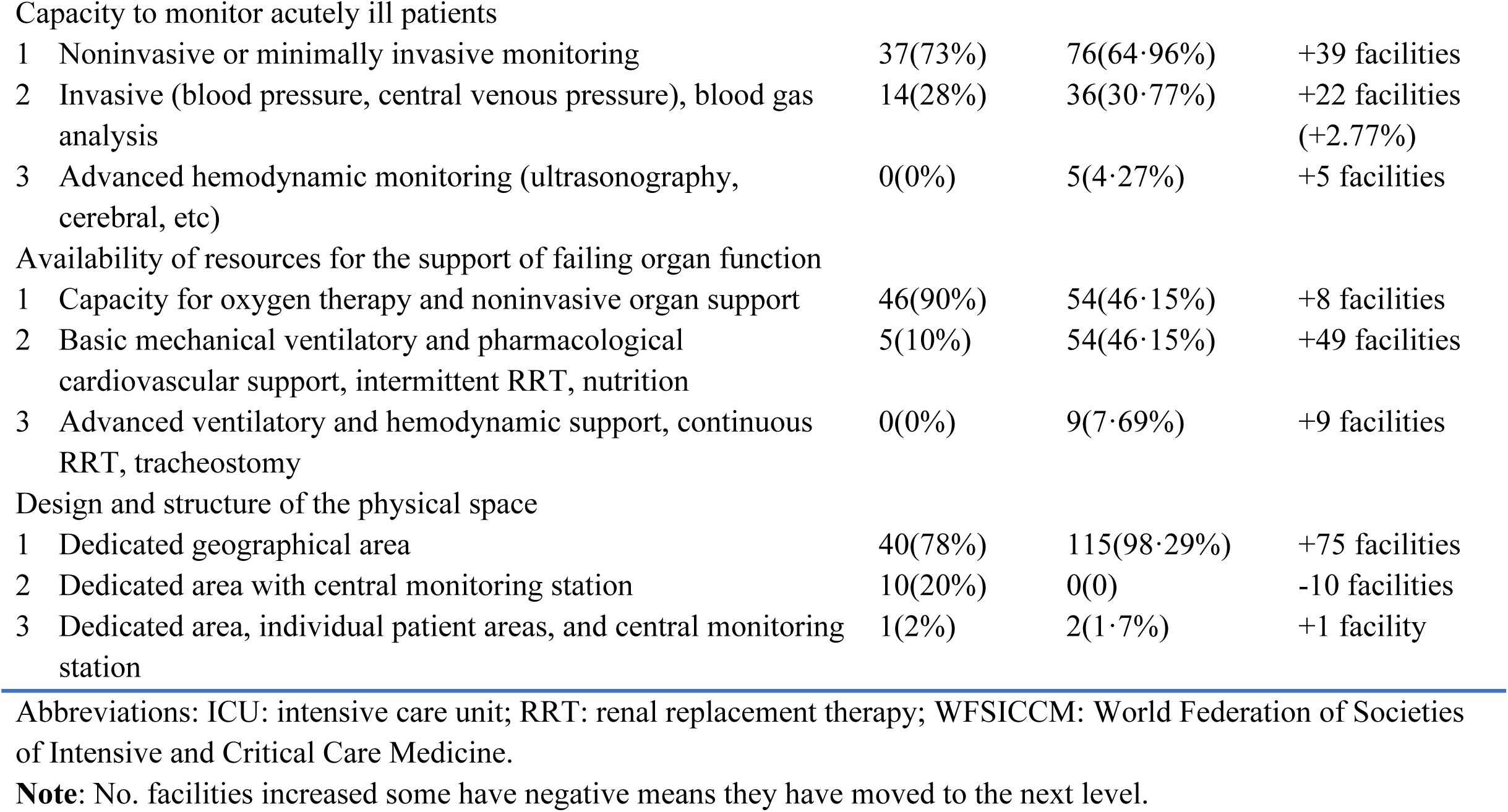
Comparison of Service Levels Pre- and Post-COVID 19 for Each of the WFSICCM Variables 1–6.

**Table 3:**
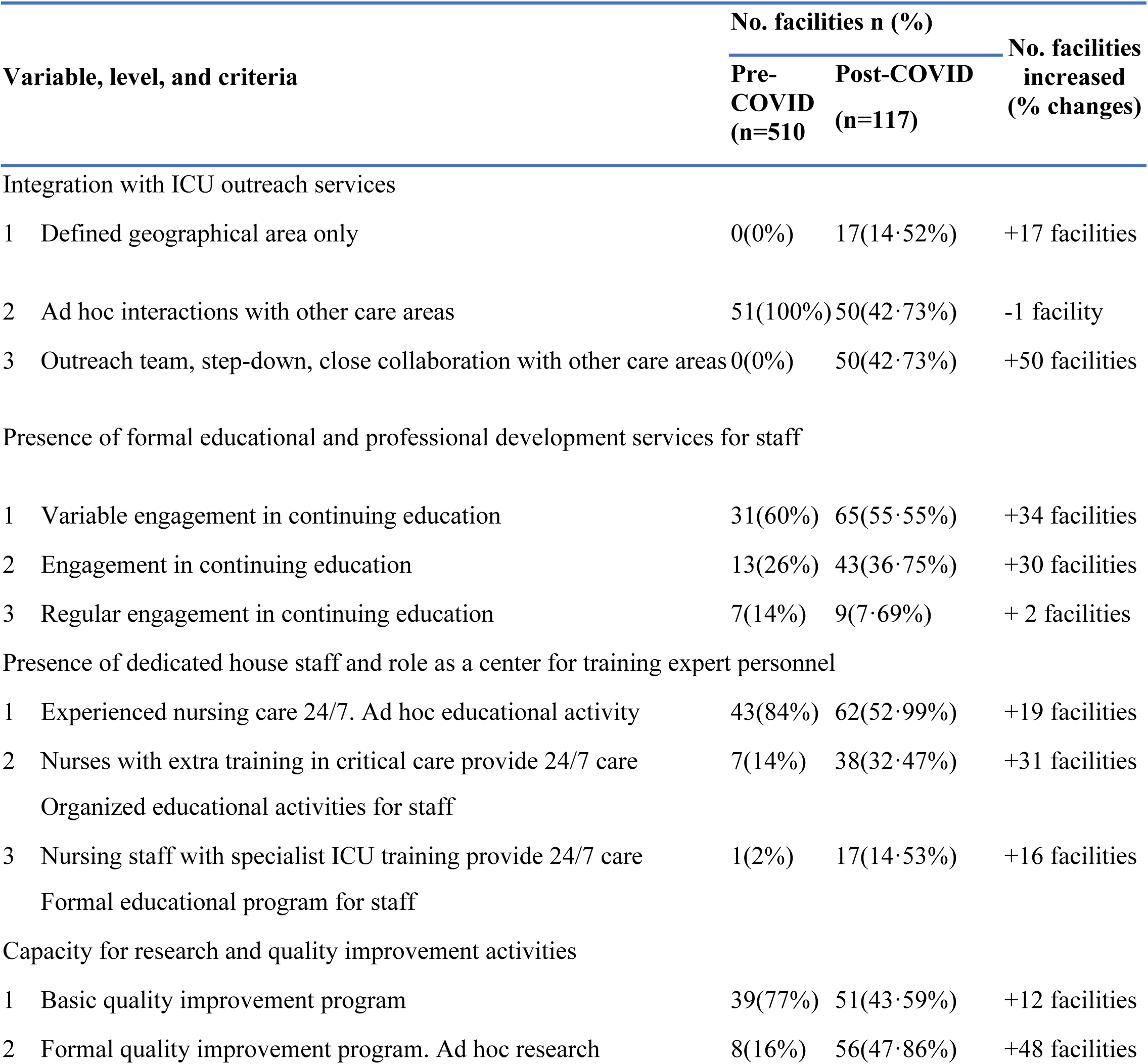
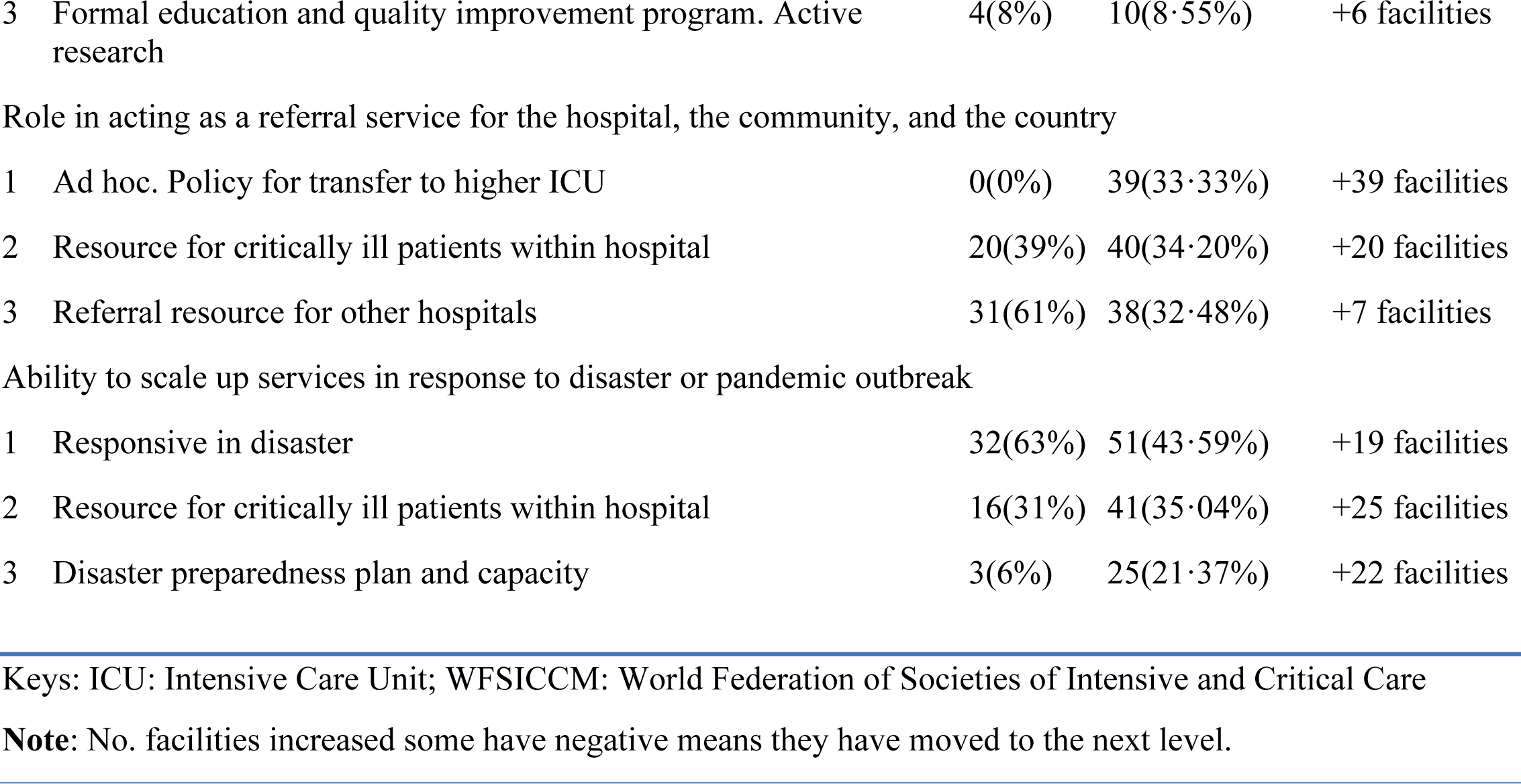
Comparison of Service Levels Pre and Post-COVID-19 for Each of the WFSICCM Variables 7– 12.

### Current Status of Private Facilities

Most private ICUs are concentrated in Addis Ababa 32/42(76·2%), where there are 21 private hospitals with ICU services, compared to 13 in the public sector. Across other regions, the presence of private ICUs is minimal, and in six regions with no private ICUs, with the majority of ICU services being provided by public facilities, the overall private bed capacity is 266/1028(25·88%) of the national bed capacity).

A high proportion of private facilities, 37/42 (88·09%) have physicians with formal ICU training available 24/7, while a significant number 27/42(64·29%), provide basic mechanical and pharmacological support for organ function. However, advanced organ support resources and hemodynamic monitoring are limited, with only 3/42(7·14%) offering such capabilities. Many private facilities show a strong dedication to education and quality improvement, with 10/42(23·81%) participating in ongoing education and 27/42(64·28%) implementing basic quality improvement programs. Only 2/42(4·76%) of facilities provide formal ICU training programs for their staff. Integration with ICU outreach services is moderate, with 24/42(57·14%) collaborating with other care areas. In 2/42(4·76%) of facilities, disaster preparedness plans are in place, and 5/42 (11·90%) of these facilities serve as referral resources for other hospitals, as further shown in **Table 4**.

**Table 4:** Current Status of Private Facilities for Each of the WFSICCM Variables 1–6.

| Variable, level, and criteria |  | No. of Private facilities n (%)<br>(n=42) |
| --- | --- | --- |
| Availability of skilled medical personnel |  |  |
| 1 | Physicians with some critical care experience at least during the day. Variable access to other specialists | 3(7.14%) |
| 2 | Physicians with ICU training/experience available during day and night. Ready access to other specialists | 2(4.76%) |
| 3 | Physicians with formal ICU training 24/7. Rapid access to full complement of specialists | 37(88.09%) |
| Nurse to patient ratio |  |  |
| 1 | Higher than ward nurse to patient ratio | 1(2.38%) |
| 2 | Not <1:3 | 17(40.47%) |
| 3 | Not <1:2 | 24(57.14%) |
| Availability of other specialists—respiratory therapists, physiotherapists, nutritionists, etc. |  |  |
| 1 | Other personnel available | 10(23.81%) |
| 2 | Variable inclusion of allied health personnel | 24(57.14%) |
| 3 | Allied health personnel as regular team members | 8(19.04%) |
| Capacity to monitor acutely ill patients |  |  |
| 1 | Noninvasive or minimally invasive monitoring | 13(30.95%) |
| 2 | Invasive (blood pressure, central venous pressure), blood gas analysis | 26(61.90%) |
| 3 | Advanced hemodynamic monitoring (ultrasonography, cerebral, etc) | 3(7.14%) |
| Availability of resources for the support of failing organ function |  |  |
| 1 | Capacity for oxygen therapy and noninvasive organ support | 12(28.57%) |
| 2 | Basic mechanical ventilatory and pharmacological cardiovascular support, intermittent RRT, nutrition | 27(64.29%) |
| 3 | Advanced ventilatory and hemodynamic support, continuous RRT, tracheostomy | 3(7.14%) |
| Design and structure of the physical space |  |  |
| 1 | Dedicated geographical area | 30(71.42%) |
| 2 | Dedicated area with central monitoring station | 3(7.14%) |
| 3 | Dedicated area, individual patient areas, and central monitoring station | 9(21.42%) |

Abbreviations: ICU: intensive care unit; RRT: renal replacement therapy; WFSICCM: World Federation of Societies of Intensive and Critical Care Medicine.

### Patient-Level Information and ICU Utilization

Among the 279 ICU admissions assessed, 218/279 (78·1%) were from the public sector and 61/279 (21·9%) from the private sector. The patients were mainly from Addis Ababa 79/279(28·3%), Oromia 64/279 (22·9%), and Amhara 45/279 (16·1%) regions. Additionally, 29 hospitals (18·2%) reported no admitted patients on the day of assessment. The mean age of patients was 39·1 years, and males accounted for 154/279 (55·2%) of all patients. Neurological conditions 87/271 (32·1%), respiratory problems 70/271 (25·8%) and sepsis 82/279(29.4%) were the main reasons for admission, with gastrointestinal and hematological conditions more common in sepsis patients. Hypertension 70/279 (25·1%) and diabetes mellitus 48/279 (17·2%) were the most frequent comorbidities, with higher rates of diabetes mellitus 17/48 (35·4%), and HIV 6/9(66·7%) in sepsis patients. Readmissions were low overall, 15/279 (5·4%), slightly higher among sepsis patients, 4/15(26·7%), as further shown in **Table 5**.

**Table 5:**
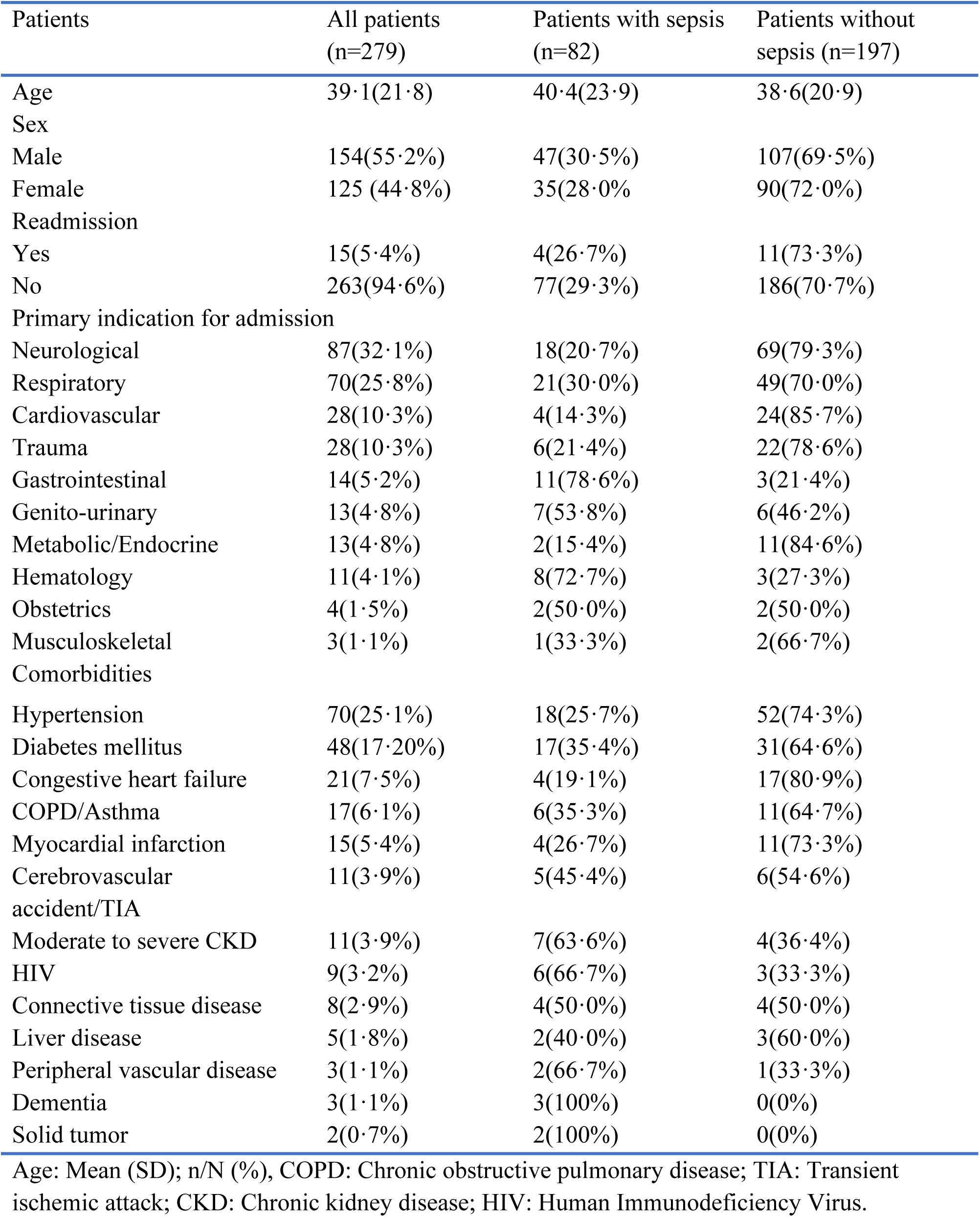
Patient level Information and ICU Utilization.

## Discussion

This nationwide assessment of ICU capacity and utilization across 159 Ethiopian hospitals representing both public and private sectors demonstrates substantial gains in critical care infrastructure and human resources over the past five years. The ICU bed capacity has tripled, increasing from 324 to 1028 beds, with the number of hospitals offering ICU services rising from 51 to 117. This corresponds to a national ICU density increase from 0.3 to 0.9 beds per 100,000 population. While this still falls short of the global average of 5-30 ICU beds per 100,000, it represents a remarkable achievement for a low-resource setting, particularly when benchmarked against regional comparators that have reported only marginal improvements.

The regional distribution of ICUs in Ethiopia has also improved post-COVID-19, with significant expansions in Addis Ababa, Oromia, Somali, Sidama, and the restructured southern regions, reflecting a shift toward decentralizing critical care. This trend aligns with findings from big African regional studies, which emphasized that regional disparities in ICU access and outcomes are common across Africa and that expanding services beyond urban hubs is critical for improving survival in critically ill patients(11,17,18). However, regions such as Benishangul-Gumuz have seen no change, highlighting persistent inequities also noted in Nigerian and Ugandan assessments, where ICU resources remain concentrated in major cities(18,19). These disparities underscore the need for targeted regional investment and support for essential emergency and critical care (EECC) in all facilities, as advocated by EECC consensus recommendations and WHO-aligned system reviews(20,21).

From a service readiness perspective, this study documented significant post-COVID-19 improvements. The availability of 24/7 ICU-trained physicians increased from 29% to 52.1%, and disaster preparedness plans rose from 6% to 21.4% of public facilities. This improvement likely stems from the MOH’s collaboration with the WHO and other stakeholders, which helped to recruit 45,000 temporary staff, and the training programs being implemented to provide timely training programs for over 2,500 clinicians, significantly strengthening the availability of a trained healthcare workforce in the ICUs (22–24).

Despite this progress, the study highlights ongoing limitations in advanced monitoring and organ support. Only 5% of facilities offer invasive hemodynamic monitoring or continuous renal replacement therapy (CRRT). These constraints are consistent with findings from both the African COVID-19 Critical Care Outcomes Study (ACCCOS) and the African Critical Illness Outcomes Study (ACIOS), which reported widespread deficits in access to advanced technologies, including vasopressors, mechanical ventilation, and renal replacement therapy across African ICUs (11,17,25). Contributing factors include prohibitive costs, limited infection control infrastructure, and a shortage of trained personnel, which continue to undermine complex critical care delivery in resource-constrained settings (26–28). While such resources are essential for managing circulatory collapse and improving outcomes in septic shock, accounting for 29% of ICU admissions in this study, they remain inaccessible to most facilities(29,30). Non-invasive technologies, such as continuous cardiac output monitoring and bedside ultrasound, have shown promise in other low-resource contexts as safer, scalable, and cost-effective alternatives, warranting implementation research and policy attention in the Ethiopian setting.

A unique strength of this study is its inclusion of private healthcare facilities, which contribute 10·8% of the nation’s ICU bed capacity. Predominantly located in Addis Ababa, these facilities generally demonstrate stronger specialist coverage and greater engagement in quality improvement initiatives compared to public institutions. However, both sectors face persistent constraints in disaster preparedness, advanced monitoring, and structured referral systems. These challenges are compounded by weak public-private integration, limited joint training opportunities, and inconsistent adherence to national referral protocols, issues echoed in broader health systems reviews across sub-Saharan Africa that emphasize the critical role of formalized public-private partnerships (PPPs) in promoting equity, efficiency, and service quality in critical care delivery (6,26,28).

Beyond capacity metrics, this study offers rare insight into ICU patient characteristics, revealing a young patient population (mean age 39.1 years), male predominance (55.2%), and leading admission causes such as neurological (32.1%), respiratory (25.8%), and sepsis (29.4%) conditions. These findings align with reports from Uganda, Tanzania, and multicountry studies like ACCCOS and ACIOS, which similarly highlight the high burden of sepsis and non-communicable complications among ICU patients in Africa (17,19,31,32). The increased readmission rate among sepsis patients shows the systemic gaps in infection prevention, antimicrobial stewardship, and post-ICU continuity of care. Importantly, the study reinforces the global call, advanced by the Essential Emergency and Critical Care (EECC) initiative and the ACCCOS findings, that life-saving interventions such as oxygen therapy, intravenous fluids, and airway management should be universally available, not restricted to high-tech ICUs (10,17,32). In Ethiopia, where many hospitals without formal ICUs still manage critically ill patients in general wards, future investment must not only focus on expanding ICU infrastructure but also on scaling up EECC across all facility levels to ensure equitable and effective care nationwide.

The main limitation of this study is that it does not assess patient outcome information, as its primary aim was to understand basic admission demographics and ICU capacity. Additionally, some facilities were excluded due to logistical challenges, which may slightly limit the comprehensiveness of the data. Despite these limitations, the study effectively addresses its primary research questions and provides a robust, comprehensive foundation for future research and sub-studies, offering valuable insights to guide improvements in critical care services and inform evidence-based interventions.

Ethiopia’s critical care landscape has significantly evolved over the past five years, with ICU capacity tripling, a more balanced regional distribution, and meaningful improvements in the deployment of human resources. The inclusion of private facilities, improved coordination efforts, and broader access to essential care reflect growing national capacity and system maturity. However, persistent limitations in monitoring, organ support, and emergency readiness point to the need for more strategic, inclusive, and technology-adaptive approaches. Anchoring future investments in EECC, public-private integration, and data-driven coordination will be essential for building a resilient, equitable critical care system capable of meeting both current and emerging health challenges.

## Data Availability

Data will be available upon reasonable request of the corresponding author.

## Contributors

All authors contributed to conceptualizing the study, writing the original draft, reviewing, and editing. Tesfay Yohannes and Fitsum K Belachew had direct access to the data, performed data linkage, data extraction, and analysis, and verified the underlying data reported. All authors have read and approved the final version of the manuscript.

## Declaration of interests

The authors declare no competing interests.

## Data sharing

Data utilized in this study will be available upon request. All shared data will be anonymized to ensure that no identifiable information is disclosed. For requests, please contact us at. There is no fixed end date for data availability.

## Acknowledgments

We extend our gratitude to the Ethiopian Ministry of Health for their collaboration and support in facilitating data collection across regions. We thank the regional health bureau focal persons, hospital administrators, and ICU staff for their cooperation during site visits. We acknowledge the dedication of the data collectors and supervisors who contributed to this nationwide survey. We are grateful to the ENCCA collaborators for their technical and logistical support.

## Funding

This research was funded by the Ministry of Health, Ethiopia and partially sponsored by the Project Network for Perioperative and Critical Care (www.n4pcc.com).

## List of supplementary files

- **Appendix A:** List of ENCCA collaborators
- **Appendix B:** Details of Private ICU facilities

## Notes

### Competing Interest Statement

The authors have declared no competing interest.

### Clinical Trial

NA

### Author Declarations

Ethical approval was obtained from the Institutional Review Board (IRB) of Hawassa University College of Medicine and Health Sciences (protocol number IRB/333/16)

